# Serum total cholesterol and fatal subarachnoid hemorrhage in 120,000 Japanese: a pooled analysis of data from 12 cohorts

**DOI:** 10.1101/2024.10.15.24315569

**Authors:** Atsushi Satoh, Hisatomi Arima, Sachiko Tanaka-Mizuno, Akira Fujiyoshi, Aya Kadota, Katsuyuki Miura, Hirotsugu Ueshima, Tomonori Okamura, Yoshitaka Murakami, EPOCH JAPAN Research Group

## Abstract

**Background:** The aim of this study was to clarify the association between serum total cholesterol and fatal subarachnoid hemorrhage in Japanese men and women.

**Methods:** The study involved a pooled analysis of data from 12 well-qualified cohort studies conducted in Japan. A total of 120,973 participants (70,947 women and 50,026 men) aged 18– 96 years at baseline underwent follow-up for a median of 12.7 years. The participants were classified according to their serum total cholesterol levels: <4.14 mmol/L (<160 mg/dL), 4.14–4.64 mmol/L (160–180 mg/dL), 4.65–5.16 mmol/L (180–199 mg/dL), 5.17–5.68 mmol/L (200–219 mg/dL), 5.69–6.20 mmol/L (220–239 mg/dL), and ≥6.21 mmol/L (≥240 mg/dL). The outcome of the analysis was death from subarachnoid hemorrhage.

**Results:** During the median follow-up of 12.7 years, 261 participants died from subarachnoid hemorrhage. In women, both low (<5.69 mmol/L [<220 mg/dL]) and high (≥6.21 mmol/L [≥240 mg/dL]) serum total cholesterol levels were significantly associated with a higher risk of fatal subarachnoid hemorrhage compared with the reference group (5.69–6.20 mmol/L [220–239 mg/dL]). These associations remained significant after adjustment for confounding factors. In contrast, no associations were observed in men.

**Conclusions:** Both low and high serum total cholesterol levels were associated with a higher risk of fatal subarachnoid hemorrhage in 70,947 female participants from 12 cohort studies throughout Japan.

## Introduction

The global incidence rate of subarachnoid hemorrhage is estimated at 9 per 100,000 person-years.^1^ However, the rate of subarachnoid hemorrhage in Japan is much higher, at 20–100 per 100,000 person-years.^1, 2^ Subarachnoid hemorrhage is the most lethal type of stroke in Japan^3^ as well as in other countries.^4, 5^ Effective prevention measures for subarachnoid hemorrhage require strategies based on reliable knowledge of risk factors for its development.

Low serum cholesterol levels have been shown to be associated with hemorrhagic stroke, including intracerebral hemorrhage and subarachnoid hemorrhage.^6-8^ However, there is no definitive evidence for a significant association between serum cholesterol and subarachnoid hemorrhage.^9-12^ The objective of the present study was to clarify the association between serum total cholesterol and fatal subarachnoid hemorrhage using data from an integrated dataset based on 12 cohort studies conducted throughout Japan.

## Methods

### Study population

The Evidence for Cardiovascular Prevention from Observational Cohorts in Japan (EPOCH-JAPAN) study was a pooled analysis of data from 15 well-qualified cohort studies conducted in Japan. The details of the project were described previously.^13^ Each cohort obtained written informed consent from the study participants and received ethical approval from the ethics committee of the relevant institute. For the present study, three cohorts in the EPOCH-JAPAN study were excluded due to a lack of data on deaths from subarachnoid hemorrhage. Consequently, 132,826 participants aged 18–96 years at baseline from 12 cohorts were included in the study. The baseline years were 1985 to 2002, and the median follow-up was 12.7 years. After exclusion of 11,853 participants with missing data on death from subarachnoid hemorrhage, serum total cholesterol, age, or sex, a total of 120,973 participants (70,947 women and 50,026 men) were included in the analysis.

### Measurements

Blood samples were collected and serum total cholesterol levels were measured using enzyme methods. Measurement of serum total cholesterol was standardized in five cohorts such that the values would be trackable to the Centers for Disease Control and Prevention reference method. The participants were divided into six groups according to their serum total cholesterol levels: <4.14 mmol/L (<160 mg/dL), 4.14–4.64 mmol/L (160–180 mg/dL), 4.65– 5.16 mmol/L (180–199 mg/dL), 5.17–5.68 mmol/L (200–219 mg/dL), 5.69–6.20 mmol/L (220–239 mg/dL), and ≥6.21 mmol/L (≥240 mg/dL). The reference group was defined as 5.69–6.20 mmol/L (220–239 mg/dL). Blood pressure was measured with a standard sphygmomanometer in all cohorts. In several cohorts where blood pressure was measured more than once, the average values were used for the analysis. Body mass index was calculated as weight in kilograms divided by height in meters squared (kg/m^2^). Smoking status and alcohol intake were measured by a questionnaire and defined as current or not.

### Outcome

For each deceased subject, the primary underlying cause of death was coded according to the International Classification of Diseases (ICD) for the National Vital Statistics based on the criteria proposed by the World Health Organization. Detailed causes of death were sought using all available sources in each cohort study. In many studies, death certificates were reviewed and/or the National Vital Statistics were utilized after obtaining permission from the Ministry of Internal Affairs and Communications of Japan. Other sources utilized in some studies included autopsy findings, medical records, health examinations, and questionnaires. The outcome of the present analysis was death from subarachnoid hemorrhage (430 in ICD-9 or I60 in ICD-10).

### Statistical analysis

For the participant characteristics, continuous variables were presented as mean and standard deviation (SD), and categorical variables were shown as number and percentage. Multivariable-adjusted hazard ratios (HRs) for fatal subarachnoid hemorrhage defined by serum total cholesterol levels were calculated using Cox proportional hazards models adjusted for age (model 1) and BMI, blood pressure, smoking, and alcohol drinking (model 2) as covariates. A Fine and Gray model analysis was conducted to account for competing risk of death from any other cause than subarachnoid hemorrhage. The significance level was set at *P*<0.05. All statistical analyses were performed using SAS version 9.4 (SAS Institute Inc., Cary, NC, USA).

## Results

The characteristics of the participants in the 12 cohorts are shown in Table 1. The mean serum total cholesterol levels ranged from 4.90 to 5.44 mmol/L (189.3 to 210.2 mg/dL). The characteristics of the female participants according to their serum total cholesterol levels are shown in Table 2. Women with higher serum total cholesterol levels were older, more likely to be obese, and less likely to be current smokers. Similar patterns were observed in men (Table 3).

**Table 1.** Characteristics of the participants in the 12 cohorts included in the present study.

| Cohort | *n* | Baseline<br>year | Women<br>*n* (%) | Age, years<br>mean (SD) | Serum TC, mmol/L<br>mean (SD) | Follow-up, years<br>median (IQR) | SAH death<br>*N* |
| --- | --- | --- | --- | --- | --- | --- | --- |
| Aichi, Workers | 5,589 | 1997 | 1,206 (21.6) | 48.1 (7.1) | 5.44 (0.91) | 10.9 (9.5–10.9) | 2 |
| CIRCS, Osaka | 6,180 | 1985 | 3,952 (63.9) | 55.1 (12.8) | 5.43 (0.95) | 13.5 (10.8–13.8) | 5 |
| Hisayama | 2,735 | 1988 | 1,573 (57.5) | 59.5 (12.1) | 5.38 (1.09) | 14.0 (14.0–14.0) | 16 |
| Iwate | 26,455 | 2002 | 17,300 (65.4) | 62.1 (11.6) | 5.18 (0.86) | 2.2 (2.0–4.1) | 13 |
| JACC | 29,008 | 1988 | 18,460 (63.6) | 57.4 ( 9.6) | 5.13 (0.95) | 18.5 (17.1–19.5) | 107 |
| JMS | 12,236 | 1992 | 7,437 (60.8) | 55.3 (11.6) | 4.97 (0.91) | 11.1 (10.3–12.0) | 23 |
| Ohasama | 2,712 | 1987 | 1,636 (60.3) | 56.2 (12.2) | 5.06 (0.95) | 14.7 (11.8–14.7) | 12 |
| Osaki | 16,044 | 1994 | 9,232 (57.5) | 62.4 (9.4) | 5.29 (0.91) | 12.7 (12.6–12.8) | 44 |
| RERF | 4,610 | 1986 | 3,105 (67.4) | 62.1 (12.3) | 5.44 (1.03) | 19.9 (12.2–20.6) | 11 |
| Suita | 6,445 | 1989 | 3,355 (52.1) | 55.4 (13.3) | 5.36 (0.96) | 14.9 (13.5–15.9) | 11 |
| Tanno-Sobetsu | 1,972 | 1977 | 1,072 (54.4) | 49.4 (8.1) | 4.90 (0.97) | 27.0 (17.1–29.4) | 13 |
| Toyama, Workers | 6,987 | 1990 | 2,619 (37.5) | 37.9 (10.0) | 4.93 (0.92) | 20.1 (19.2–20.7) | 4 |

TC: total cholesterol; SAH: subarachnoid hemorrhage; SD: standard deviation; IQR: interquartile range; CIRCS: Circulatory Risk in Communities Study; JACC: Japan Collaborative Cohort Study; JMS: Jichi Medical School; RERF: Radiation Effects Research Foundation.

**Table 2.** Baseline characteristics of the female participants according to their serum total cholesterol levels.

|  | Serum total cholesterol, mmol/L |  |  |  |  |  | *P*<br>value |
| --- | --- | --- | --- | --- | --- | --- | --- |
|  | <4.14<br>(<160 mg/dL) | 4.14–4.64<br>(160–179<br>mg/dL) | 4.65–5.16<br>(180–199<br>mg/dL) | 5.17–5.68<br>(200–219<br>mg/dL) | 5.69–6.20<br>(220–239<br>mg/dL) | ≥6.21<br>(≥240 mg/dL) |  |
| Number of participants | 6,496 | 10,234 | 15,118 | 15,526 | 11,676 | 11,717 |  |
| Age, years | 50.7 (14.1) | 54.0 (13.2) | 56.8 (12.3) | 58.9 (11.0) | 60.1 (10.2) | 60.5 (9.4) | <0.001 |
| BMI, kg/m<sup>2</sup> | 22.4 (3.2) | 22.9 (3.3) | 23.3 (3.4) | 23.6 (3.3) | 23.8 (3.3) | 24.1 (3.4) | <0.001 |
| History of CVD, *n* (%) | 234 (3.7) | 379 (3.7) | 673 (4.5) | 666 (4.3) | 573 (4.9) | 632 (5.4) | <0.001 |
| Smoking, *n* (%) |  |  |  |  |  |  |  |
| Current smokers | 395 (6.4) | 541 (5.6) | 672 (4.7) | 600 (4.1) | 503 (4.6) | 531 (4.8) | <0.001 |
| Previous smokers | 154 (2.5) | 192 (2.0) | 252 (1.8) | 238 (1.6) | 189 (1.7) | 233 (2.1) |  |
| Alcohol drinking, *n* (%) |  |  |  |  |  |  |  |
| Current drinkers | 1,515 (24.9) | 2,322 (24.0) | 3,187 (22.2) | 3,093 (21.1) | 2,228 (20.3) | 2,135 (19.3) | <0.001 |
| Previous drinkers | 92 (1.5) | 147 (1.5) | 195 (1.4) | 244 (1.7) | 183 (1.7) | 181 (1.6) |  |
| Total cholesterol, mmol/L | 3.78 (0.30) | 4.41 (0.15) | 4.91 (0.15) | 5.41 (0.15) | 5.92 (0.15) | 6.78 (0.57) | <0.001 |
| Systolic BP, mmHg | 121.7 (19.4) | 124.9 (19.6) | 127.3 (19.8) | 129.5 (20.1) | 131.2 (20.4) | 133.6 (20.3) | <0.001 |
| Diastolic BP, mmHg | 72.1 (11.5) | 74.2 (11.5) | 75.6 (11.4) | 76.8 (11.4) | 77.6 (11.6) | 79.1 (11.4) | <0.001 |

BMI: body mass index; CVD: cardiovascular disease; BP: blood pressure.

**Table 3.**
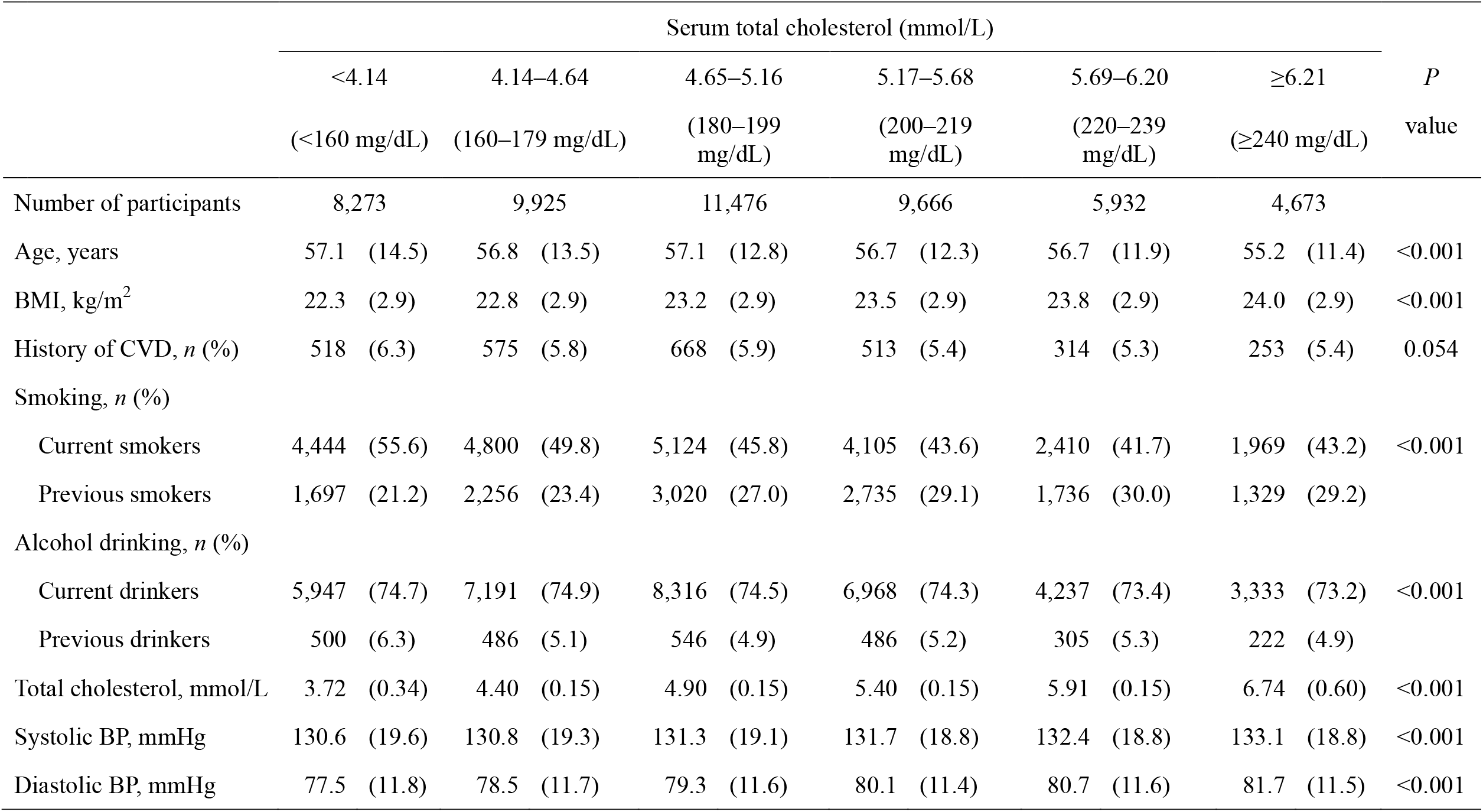
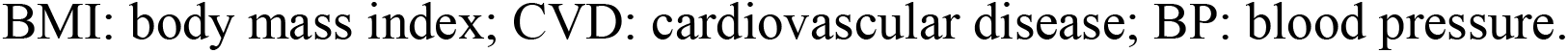
Baseline characteristics of the male participants according to their serum total cholesterol levels.

|  | Serum total cholesterol (mmol/L) |  |  |  |  |  | *P*<br>value |
| --- | --- | --- | --- | --- | --- | --- | --- |
|  | <4.14<br>(<160 mg/dL) | 4.14–4.64<br>(160–179 mg/dL) | 4.65–5.16<br>(180–199 mg/dL) | 5.17–5.68<br>(200–219 mg/dL) | 5.69–6.20<br>(220–239 mg/dL) | ≥6.21<br>(≥240 mg/dL) |  |
| Number of participants | 8,273 | 9,925 | 11,476 | 9,666 | 5,932 | 4,673 |  |
| Age, years | 57.1 (14.5) | 56.8 (13.5) | 57.1 (12.8) | 56.7 (12.3) | 56.7 (11.9) | 55.2 (11.4) | <0.001 |
| BMI, kg/m<sup>2</sup> | 22.3 (2.9) | 22.8 (2.9) | 23.2 (2.9) | 23.5 (2.9) | 23.8 (2.9) | 24.0 (2.9) | <0.001 |
| History of CVD, *n* (%) | 518 (6.3) | 575 (5.8) | 668 (5.9) | 513 (5.4) | 314 (5.3) | 253 (5.4) | 0.054 |
| Smoking, *n* (%) |  |  |  |  |  |  |  |
| Current smokers | 4,444 (55.6) | 4,800 (49.8) | 5,124 (45.8) | 4,105 (43.6) | 2,410 (41.7) | 1,969 (43.2) | <0.001 |
| Previous smokers | 1,697 (21.2) | 2,256 (23.4) | 3,020 (27.0) | 2,735 (29.1) | 1,736 (30.0) | 1,329 (29.2) |  |
| Alcohol drinking, *n* (%) |  |  |  |  |  |  |  |
| Current drinkers | 5,947 (74.7) | 7,191 (74.9) | 8,316 (74.5) | 6,968 (74.3) | 4,237 (73.4) | 3,333 (73.2) | <0.001 |
| Previous drinkers | 500 (6.3) | 486 (5.1) | 546 (4.9) | 486 (5.2) | 305 (5.3) | 222 (4.9) |  |
| Total cholesterol, mmol/L | 3.72 (0.34) | 4.40 (0.15) | 4.90 (0.15) | 5.40 (0.15) | 5.91 (0.15) | 6.74 (0.60) | <0.001 |
| Systolic BP, mmHg | 130.6 (19.6) | 130.8 (19.3) | 131.3 (19.1) | 131.7 (18.8) | 132.4 (18.8) | 133.1 (18.8) | <0.001 |
| Diastolic BP, mmHg | 77.5 (11.8) | 78.5 (11.7) | 79.3 (11.6) | 80.1 (11.4) | 80.7 (11.6) | 81.7 (11.5) | <0.001 |

BMI: body mass index; CVD: cardiovascular disease; BP: blood pressure.

During the median follow-up of 12.7 years, 261 participants died from subarachnoid hemorrhage. The associations between serum total cholesterol and fatal subarachnoid hemorrhage in women and men are shown in Table 4. In women, both high and low serum total cholesterol levels were significantly associated with a higher risk of fatal subarachnoid hemorrhage compared with the reference group. These associations remained significant in the multivariable analysis with adjustment for body mass index, systolic blood pressure, smoking, and alcohol drinking compared with the reference group. Comparable findings were observed in the Fine and Gray model accounting for competing risk of death from any other cause than subarachnoid hemorrhage; the HR (95% confidence interval) values were 2.73 (1.34–5.55) for serum total cholesterol <4.14 mmol/L (<160 mg/dL), 1.97 (1.01–3.83) for serum total cholesterol 4.14–4.64 mmol/L (160–180 mg/dL), 2.01 (1.09–3.70) for serum total cholesterol 4.65–5.16 mmol/L (180–199 mg/dL), 1.95 (1.06–3.56) for serum total cholesterol 5.17 to 5.68 mmol/L (200–219 mg/dL), and 2.17 (1.18–3.97) for serum total cholesterol ≥6.21 mmol/L (≥240 mg/dL) compared with the reference group. In contrast, there were no significant associations between serum total cholesterol and fatal subarachnoid hemorrhage in men. We also conducted analyses in women stratified by age (65 and 55 years), as shown in Supplemental Tables 1 and 2. Significant positive associations were found for the lowest serum total cholesterol group in women aged <65 years and the highest serum total cholesterol group in women aged ≥65 years. In women aged ≥55 years, all serum total cholesterol groups had a significantly higher risk of fatal subarachnoid hemorrhage compared with the reference group, while no association was found in women aged <55 years. Similar associations were found in the analysis excluding participants with history of cardiovascular disease (Supplemental Table 3).

**Table 4.** Associations between serum total cholesterol and fatal subarachnoid hemorrhage in women and men.

|  |  | Serum total cholesterol, mmol/L |  |  |  |  |  |
| --- | --- | --- | --- | --- | --- | --- | --- |
|  |  | <4.14<br>(<160 mg/dL) | 4.14–4.64<br>(160–179 mg/dL) | 4.65–5.16<br>(180–199 mg/dL) | 5.17–5.68<br>(200–219 mg/dL) | 5.69–6.20<br>(220–239 mg/dL) | ≥6.21<br>(≥240 mg/dL) |
| Women |  |  |  |  |  |  |  |
| Number of participants |  | 6,496 | 10,234 | 15,118 | 15,526 | 11,676 | 11,717 |
| Number of SAH deaths |  | 17 | 22 | 41 | 40 | 19 | 41 |
| Hazard ratio (95% CI) |  |  |  |  |  |  |  |
| Model 1 |  | 2.31<br>(1.19–4.47) | 1.64<br>(0.88–3.03) | 1.91<br>(1.11–3.29) | 1.68<br>(0.98–2.92) | Reference | 2.00<br>(1.16–3.45) |
| Model 2 |  | 3.05<br>(1.49–6.25) | 2.07<br>(1.06–4.03) | 2.08<br>(1.13–3.82) | 1.96<br>(1.07–3.59) | Reference | 2.14<br>(1.17–3.92) |
| Men |  |  |  |  |  |  |  |
| Number of participants |  | 8,273 | 9,925 | 11,476 | 9,666 | 5,932 | 4,673 |
| Number of SAH deaths |  | 24 | 19 | 14 | 14 | 7 | 3 |
| Hazard ratio (95% CI) |  |  |  |  |  |  |  |
| Model 1 |  | 2.17<br>(0.93–5.08) | 1.50<br>(0.63–3.59) | 0.98<br>(0.39–2.43) | 1.19<br>(0.48–2.95) | Reference | 0.53<br>(0.14–2.06) |
| Model 2 |  | 1.65<br>(0.69–3.93) | 1.20<br>(0.49–2.91) | 0.83<br>(0.33–2.09) | 1.16<br>(0.47–2.89) | Reference | 0.54<br>(0.14–2.10) |

SAH: subarachnoid hemorrhage; CI: confidence interval. Model 1: adjusted for age. Model 2: adjusted for age, body mass index, systolic blood pressure, smoking, and alcohol drinking.

## Discussion

In the present pooled analysis using data from the integrated EPOCH-JAPAN dataset covering 12 cohort studies throughout Japan, both high (≥6.21 mmol/L [≥240 mg/dL]) and low (<5.69 mmol/L [<220 mg/dL]) serum total cholesterol levels in women were associated with a higher risk of fatal subarachnoid hemorrhage compared with the reference group (5.69–6.20 mmol/L [220–239 mg/dL]). These associations remained significant after adjustment for confounding factors such as body mass index, systolic blood pressure, smoking, and alcohol drinking. Low cholesterol was associated with the risk of fatal subarachnoid hemorrhage in women aged ≤65 years, whereas high cholesterol was associated with the risk of fatal subarachnoid hemorrhage in women aged >65 years. In contrast, no associations between serum total cholesterol and fatal subarachnoid hemorrhage were noted in men, although a non-significant trend toward an increased risk of fatal subarachnoid hemorrhage was observed for very low serum total cholesterol levels (<4.14 mmol/L [160 mg/dL]).

Several prospective studies have investigated the association between serum total cholesterol and incidence or mortality of subarachnoid hemorrhage, but no studies have examined the association between serum total cholesterol and risk of subarachnoid hemorrhage separately for men and women. The Honolulu Heart Program found no clear association between serum total cholesterol and incidence of subarachnoid hemorrhage in Japanese American populations,^9^ while the JACC study found no association between serum total cholesterol and mortality from subarachnoid hemorrhage in Japanese subjects.^11^

Furthermore, the Asia-Pacific Cohort Studies Collaboration, a meta-analysis of data for participants in prospective cohort studies carried out in the Asia-Pacific region, revealed no clear association between serum total cholesterol and fatal/nonfatal subarachnoid hemorrhage.^10^ However, a stroke registry study conducted in Akita Prefecture, Japan, suggested an increased incidence of subarachnoid hemorrhage in Japanese people with low serum total cholesterol levels (<4.14 mmol/L [160 mg/dL]).^2^ A recent systematic review of prospective studies, including the Honolulu Heart Program and JACC study, found that low total serum cholesterol levels were associated with an increased risk of subarachnoid hemorrhage in East Asian populations, while no such associations were observed in non-East Asian populations.^14^ In the present study, low serum total cholesterol levels (<5.69 mmol/L [220 mg/dL]) were associated with an increased risk of fatal subarachnoid hemorrhage in women. In addition, very low serum total cholesterol levels (<4.14 mmol/L [160 mg/dL]) showed a tendency to be associated with an increased risk of fatal subarachnoid hemorrhage in men. Based on the above findings, low serum total cholesterol levels have the potential to be associated with an increased risk of subarachnoid hemorrhage in East Asian people.

In this pooled analysis of data from prospective studies, high serum total cholesterol levels (≥6.21 mmol/L [≥240 mg/dL]) were associated with increased mortality from subarachnoid hemorrhage in women aged ≥65 years, but no such association was observed in men. Most previous observational studies^2, 9-11^ also found no increased risk of subarachnoid hemorrhage associated with high serum total cholesterol levels. A meta-analysis revealed that postmenopausal state was associated with increased serum total cholesterol levels and higher risk of subarachnoid hemorrhage.^15^ However, data on postmenopausal status were not available for the women evaluated in the present study. Since most Japanese women aged ≥55 years would be postmenopausal,^16^ we conducted an analysis in women stratified by age of 55 years, and found a stronger association in women aged ≥55 years compared with women aged <55 years. These findings were consistent with those in a previous study.^15^ Further investigations are required to confirm whether there is a causal relationship between high serum total cholesterol and increased risk of fatal subarachnoid hemorrhage.

The mechanisms underlying the association between low serum total cholesterol and fatal subarachnoid hemorrhage, especially in women aged <65 years, remain unclear. A possible mechanism involves the promotion of arterial medial layer smooth muscle cell necrosis associated low serum total cholesterol.^17^ Low serum total cholesterol was also shown to be associated with impaired endothelium in arteries.^18^ Endothelium and smooth muscle cell impairment in the walls of cerebral artery aneurysms may promote rupture of the aneurysms and subsequent hemorrhage in the subarachnoid space.

The strengths of the present study are the very large sample size of 120,973 participants with a single ethnicity (Japanese) based on a pooled analysis of data from prospective studies and the long-term follow-up period. Several limitations of the study should also be discussed.

Because the pooled analysis included participants of community health examinations, who were more likely to be health-conscious, the findings may have been affected by selection bias. The non-standardized measurement methods for serum total cholesterol levels in some cohorts may also have resulted in misclassification of exposure. Some cohorts started earlier than the tradition for collecting data on treatment for dyslipidemia. Thus, medication for dyslipidemia was not considered in the study.

In conclusion, both low and high serum total cholesterol levels were associated with a higher risk of fatal subarachnoid hemorrhage in 70,947 women from 12 cohort studies conducted throughout Japan.

## Data Availability

The datasets used in the analysis are not available for readers.

## Acknowledgments

The Evidence for Cardiovascular Prevention from Observational Cohorts in Japan (EPOCH– JAPAN) Research Group is composed of the following investigators. Co-Chairpersons: Hirotsugu Ueshima (Shiga University of Medical Science), Tomonori Okamura (Keio University School of Medicine), and Yoshitaka Murakami (Toho University). Executive Committee: Hirotsugu Ueshima (Shiga University of Medical Science), Yutaka Kiyohara (Hisayama Research Institute for Lifestyle Diseases), Toshiharu Ninomiya (Kyushu University Graduate School of Medicine), Yutaka Imai (Tohoku Institute for Management of Blood Pressure), Takayoshi Ohkubo (Teikyo University School of Medicine), Hiroyasu Iso (The National Center for Global Health and Medicine), Isao Muraki (Osaka University Graduate School of Medicine), Kazumasa Yamagishi (University of Tsukuba Institute of Medicine), Akiko Tamakoshi (Hokkaido University Faculty of Medicine), Yoshihiro Miyamoto (National Cerebral and Cardiovascular Center), Yoshihiro Kokubo (National Cerebral and Cardiovascular Center), Katsuyuki Miura (Shiga University of Medical Science), Sachiko Tanaka-Mizuno (Kyoto University Graduate School of Medicine and Public Health), Akiko Harada (Shiga University of Medical Science), Shigeyuki Saitoh (Sapporo Medical University), Hirofumi Ohnishi (Sapporo Medical University), Ichiro Tsuji (Tohoku University Graduate School of Medicine), Atsushi Hozawa (Tohoku University), Hideaki Nakagawa (Kanazawa Medical University), Masaru Sakurai (Kanazawa Medical University), Michiko Yamada (Radiation Effects Research Foundation), Yoshimi Tatsukawa (Radiation Effects Research Foundation), Kiyomi Sakata (Iwate Medical University), Kozo Tanno (Iwate Medical University), Akihiko Kitamura (Yao City Public Health Center), Masahiko Kiyama (Institute of Preventive Medicine Inc., Osaka Center for Cancer and Cardiovascular Disease Prevention), Yuji Shimizu (Osaka Institute of Public Health), Akira Okayama (Research Institute of Strategy for Prevention), Shizukiyo Ishikawa (Jichi Medical University), Hiroshi Yatsuya (Nagoya University Graduate School of Medicine), Takeo Nakayama (Kyoto University School of Public Health), Fujiko Irie (Ibaraki Prefecture), and Toshimi Sairenchi (Dokkyo Medical University).

## Information about previous presentations

This work has not been presented previously.

## References

1. de Rooij NK, Linn FH, van der Plas JA, Algra A, Rinkel GJ. Incidence of subarachnoid haemorrhage: A systematic review with emphasis on region, age, gender and time trends. J Neurol Neurosurg Psychiatry. 2007;78:1365–1372

2. Suzuki K, Izumi M, Sakamoto T, Hayashi M. Blood pressure and total cholesterol level are critical risks especially for hemorrhagic stroke in akita, japan. Cerebrovasc Dis. 2011;31:100–106

3. Takashima N, Arima H, Kita Y, Fujii T, Miyamatsu N, Komori M, et al. Incidence, management and short-term outcome of stroke in a general population of 1.4 million japaneseshiga stroke registry. Circ J. 2017;81:1636–1646

4. Feigin VL, Lawes CMM, Bennett DA, Barker-Collo SL, Parag V. Worldwide stroke incidence and early case fatality reported in 56 population-based studies: A systematic review. The Lancet Neurology. 2009;8:355–369

5. Etminan N, Chang HS, Hackenberg K, de Rooij NK, Vergouwen MDI, Rinkel GJE, et al. Worldwide incidence of aneurysmal subarachnoid hemorrhage according to region, time period, blood pressure, and smoking prevalence in the population: A systematic review and meta-analysis. JAMA Neurol. 2019;76:588–597

6. Wang X, Dong Y, Qi X, Huang C, Hou L. Cholesterol levels and risk of hemorrhagic stroke: A systematic review and meta-analysis. Stroke. 2013;44:1833–1839

7. Ueshima H, Iida M, Shimamoto T, Konishi M, Tsujioka K, Tanigaki M, et al. Multivariate analysis of risk factors for stroke. Eight-year follow-up study of farming villages in akita, japan. Preventive medicine. 1980;9:722–740

8. Iso H, Jacobs DR, Wentworth D, Neaton JD, Cohen JD. Serum cholesterol levels and six-year mortality from stroke in 350,977 men screened for the multiple risk factor intervention trial. New England Journal of Medicine. 1989;320:904–910

9. Yano K, Reed DM, MacLean CJ. Serum cholesterol and hemorrhagic stroke in the honolulu heart program. Stroke. 1989;20:1460–1465

10. Feigin V, Parag V, Lawes CM, Rodgers A, Suh I, Woodward M, et al. Smoking and elevated blood pressure are the most important risk factors for subarachnoid hemorrhage in the asia-pacific region: An overview of 26 cohorts involving 306,620 participants. Stroke. 2005;36:1360–1365

11. Cui R, Iso H, Toyoshima H, Date C, Yamamoto A, Kikuchi S, et al. Serum total cholesterol levels and risk of mortality from stroke and coronary heart disease in japanese: The jacc study. Atherosclerosis. 2007;194:415–420

12. Sundstrom J, Soderholm M, Soderberg S, Alfredsson L, Andersson M, Bellocco R, et al. Risk factors for subarachnoid haemorrhage: A nationwide cohort of 950 000 adults. Int J Epidemiol. 2019;48:2018–2025

13. Murakami Y, Hozawa A, Okamura T, Ueshima H, Evidence for Cardiovascular Prevention From Observational Cohorts in Japan Research G. Relation of blood pressure and allcause mortality in 180,000 japanese participants: Pooled analysis of 13 cohort studies. Hypertension. 2008;51:1483–1491

14. Xie L, Wu W, Chen J, Tu J, Zhou J, Qi X, et al. Cholesterol levels and hemorrhagic stroke risk in east asian versus non-east asian populations: A systematic review and metaanalysis. The neurologist. 2017;22:107–115

15. Algra AM, Klijn CJ, Helmerhorst FM, Algra A, Rinkel GJ. Female risk factors for subarachnoid hemorrhage: A systematic review. Neurology. 2012;79:1230–1236

16. Sakata R, Shimizu Y, Soda M, Yamada M, Hsu WL, Hayashi M, et al. Effect of radiation on age at menopause among atomic bomb survivors. Radiat Res. 2011;176:787–795

17. Tirschwell DL, Smith NL, Heckbert SR, Lemaitre RN, Longstreth WT, Jr., Psaty BM. Association of cholesterol with stroke risk varies in stroke subtypes and patient subgroups. Neurology. 2004;63:1868–1875

18. Konishi M, Iso H, Komachi Y, Iida M, Shimamoto T, Jacobs DR, Jr., et al. Associations of serum total cholesterol, different types of stroke, and stenosis distribution of cerebral arteries. The akita pathology study. Stroke. 1993;24:954–964

